# Risk of Cardiovascular Events after Covid-19: a double-cohort study

**DOI:** 10.1101/2021.12.27.21268448

**Authors:** Larisa G. Tereshchenko, Adam Bishop, Nora Fisher-Campbell, Jacqueline Levene, Craig C. Morris, Hetal Patel, Erynn Beeson, Jessica A. Blank, JG N. Bradner, Michelle Coblens, Jacob W. Corpron, Jenna M. Davison, Kathleen Denny, Mary S. Earp, Simeon Florea, Howard Freeman, Olivia Fuson, Florian Guillot, Kazi Haq, Jessica Hyde, Ayesha Khader, Clinton Kolseth, Morris Kim, Olivia Krol, Lisa Lin, Liat Litwin, Aneeq Malik, Evan Mitchell, Aman Mohapatra, Cassandra Mullen, Chad D Nix, Ayodele Oyeyemi, Christine Rutlen, Lisa Corley-Stampke, Ashley Tam, Inga Van Buren, Jessica Wallace, Akram Khan

## Abstract

**Objective:** To determine absolute and relative risks of either symptomatic or asymptomatic SARS-CoV-2 infection for late cardiovascular events and all-cause mortality.

**Methods:** We conducted a retrospective double-cohort study of patients with either symptomatic or asymptomatic SARS-CoV-2 infection [COVID-19(+) cohort] and its documented absence [COVID-19(-) cohort]. The study investigators drew a simple random sample of records from all Oregon Health & Science University (OHSU) Healthcare patients (N=65,585) with available COVID-19 test results, performed 03.01.2020 - 09.13.2020. Exclusion criteria were age < 18y and no established OHSU care. The primary outcome was a composite of cardiovascular morbidity and mortality. All-cause mortality was the secondary outcome.

**Results:** The study population included 1355 patients (mean age 48.7±20.5 y; 770(57%) female, 977(72%) white non-Hispanic; 1072(79%) insured; 563(42%) with cardiovascular disease (CVD) history). During a median 6 months at risk, the primary composite outcome was observed in 38/319 (12%) COVID-19(+) and 65/1036 (6%) COVID-19(-) patients (*p*=0.001). In Cox regression adjusted for demographics, health insurance, and reason for COVID-19 testing, SARS-CoV-2 infection was associated with the risk of the primary composite outcome (HR 1.71; 95%CI 1.06-2.78; *p*=0.029). Inverse-probability-weighted estimation, conditioned for 31 covariates, showed that for every COVID-19(+) patient, the average time to all-cause death was 65.5 days less than when all these patients were COVID-19(-): average treatment effect on the treated -65.5 (95%CI -125.4 to -5.61) days; *p*=0.032.

**Conclusions:** Either symptomatic or asymptomatic SARS-CoV-2 infection is associated with increased risk of late cardiovascular outcomes and has causal effect on all-cause mortality in a late post-COVID-19 period.

ClinicalTrials.gov Identifier: NCT04555187

**Key messages:** *What is already known about this subject:*

- Acute, symptomatic COVID-19 can cause acute cardiovascular manifestations.
- Post-acute or “long” COVID-19 can be a debilitating disease following acute infection with a heterogenous presentation.

*What might this study add?:*

- Either symptomatic or asymptomatic SARS-CoV-2 infection is associated with increased risk of late cardiovascular outcomes.
- Either symptomatic or asymptomatic SARS-CoV-2 infection has causal effect on all-cause mortality in a late post-COVID-19 period.

*How might this impact on clinical practice?:*

- As we begin to care for more survivors of COVID-19, we will need to better understand not only how to care for their acute symptoms and complications following infection, but also recognize future cardiovascular risk and mitigate such risk with appropriate screening and preventative measures.

## Introduction

Severe acute respiratory syndrome coronavirus-2 (SARS-CoV-2) uses angiotensin-converting enzyme 2 (ACE2) as the receptor-binding domain.[1] Cardiac myocytes express ACE2, which makes the heart a target organ in the novel coronavirus disease 2019 (COVID-19).[2] The COVID-19 pandemic has revealed heterogenous cardiovascular manifestations of infection, which likely contribute to the high case fatality rate in COVID-19.[3] Myocardial injury in COVID-19 can be caused by both direct injury to cardiac myocytes as well as secondary effects from the systemic inflammation and hypercoagulable state seen in acute infection. Cardiovascular manifestations of COVID-19 include acute myocardial infarction, stress cardiomyopathy, myocarditis, heart failure (HF), pulmonary embolism, and cardiac arrhythmias.[3] Furthermore, COVID-19 patients with known cardiovascular disease (CVD) and other risk factors including age, hypertension, diabetes, obesity, kidney disease, and respiratory system disease are more likely to require critical care and have a higher mortality rate.[4]

Post-acute or “long” COVID-19 has been described in patients with persistent symptoms or complications after the end of the acute phase of infection.[5, 6] The virulence and transmissibility of the SARS-CoV-2 virus and ongoing difficulties with public health policies and compliance challenges the ability to control COVID-19. As the virus mutates, it continues circulating throughout the globe.[7] Acute COVID-19 cardiovascular manifestations have been described in great depth.[8, 9, 10] However, the impact of COVID-19 on long-term cardiovascular outcomes remains unknown.[10] Furthermore, previous studies have had limitations, such as selection bias and absence of a control. Thus, it is essential to study the incidence, manifestations, and risk factors of post-acute outcomes that either symptomatic or asymptomatic SARS-CoV-2 infection poses on cardiovascular health.

The COVID-19 pandemic disrupted the delivery of standard cardiovascular care[11] which led to increased cardiovascular mortality in populations presumably unexposed to the SARS-CoV-2 virus.[12, 13] On the other hand, SARS-CoV-2 infection can be asymptomatic, and, thus, undiagnosed without testing. It remains unknown if the SARS-CoV-2 infection (either symptomatic or asymptomatic) is associated with a risk of long-term cardiovascular events, as compared to verified absence of the SARS-CoV-2 infection. Furthermore, while higher than expected all-cause mortality during the pandemic has been recognized,[14] it is unclear whether either asymptomatic or symptomatic SARS-CoV-2 infections may have played a causal role. To address these knowledge gaps, we prospectively designed and conducted a retrospective double-cohort study to determine: (1) absolute (attributable) risk, (2) relative conditional risk, and (3) causal inference effect of either symptomatic or asymptomatic SARS-CoV-2 infection on post-acute (late) cardiovascular events and all-cause mortality.

## Methods

Patients or the public were not involved in the design, conduct, or reporting of this study or the study results dissemination plans. We conducted a prospectively designed retrospective double-cohort study at the Oregon Health & Science University (OHSU) in Portland, Oregon. The study has been approved by the OHSU Institutional Review Board (IRB) and was registered (http://www.clinicaltrials.gov. Unique identifier: NCT04555187). Study results are reported using STROBE guidelines; a checklist is provided as a Supplement.

### Eligibility criteria

Electronic medical records (EMRs) of adult (age ≥ 18 y) patients were eligible for inclusion in the study if there was a positive or negative COVID-19 test result available. We excluded records of children (< 18 y) and those without evidence of established medical care at OHSU. The study investigators drew a simple random sample of records from the pool of all EMRs with available results of the COVID-19 test, performed at OHSU Healthcare between March 1^st^, 2020, and September 13^th^, 2020. OHSU Healthcare included all OHSU inpatient and outpatient clinical sites, including OHSU Hospital, Hillsboro Medical Center (previously Tuality Healthcare), and Adventist Medical Center.

### Exposure: definition of a Covid19 episode

A COVID-19 episode was defined as the documented by the polymerase chain reaction (PCR) test presence [COVID-19(+)] or absence [COVID-19(-)] of SARS-CoV-2 infection. Per the study design, participants were determined to have one or two COVID-19 episodes over the study time period. The following combinations were considered for any given medical record: (1) one COVID-19(+) episode, (2) one COVID-19(-) episode, (3) first COVID-19(+) episode and second COVID-19(-) episode, (4) first COVID-19(-) episode and second COVID-19(+) episode, (5) first COVID-19(+) episode and second COVID-19(+) episode, (6) first COVID-19(-) episode and second COVID-19(-) episode. The date of each episode’s onset was defined as the date of PCR test specimen collection.

The first COVID-19(+) episode or a single COVID-19 (+) episode was defined as a positive SARS-CoV-2 test. For the data collection purpose, we assumed a 30-day length for one COVID-19(+) episode. If there were multiple PCR tests, negative PCR tests within the previous 13 days were permitted before the first COVID-19(+) episode.

All of the following conditions were required for a second COVID-19(+) episode after the first COVID-19(+) episode: (1) must be confirmed by PCR and not an antibody test; (2) occurred > 30 days after the first COVID-19(+) episode; (3) if the first COVID-19(+) episode was symptomatic, the second episode could not start until after a symptom–free period of > 7 days.

The 2^nd^ COVID-19(-) episode after the 1^st^ COVID-19(+) episode was defined if it was confirmed by PCR and occurred > 30 days after the 1^st^ PCR-confirmed COVID-19(+) episode.

The first COVID-19(-) episode or a single COVID-19(-) episode was defined as any negative SARS-CoV-2 test if there was no positive SARS-CoV-2 PCR test ≥ 14 days after. The length of the COVID-19(-) episode was assumed to be ≥ 14 days, starting on the date of the negative PCR test specimen collection. If the negative SARS-CoV-2 test was an antibody test, we assumed that the COVID-19(-) episode started on March 1^st^, 2020, and ended on the date of the negative antibody test.

The 2^nd^ COVID-19(+) after 1^st^ COVID-19(-) episode was defined if a positive PCR test occurred either (1) ≥ 14 days after a negative PCR test(s), or (2) ≥ 1 day after a negative antibody test that had defined the 1^st^ COVID-19(-) episode. A second COVID-19(-) episode after a 1^st^ COVID-19(-) episode was defined as a negative PCR SARS-CoV-2 test that occurred ≥ 14 days after the 1^st^ PCR-confirmed COVID-19(-) episode.

### Retrospective follow-up and the study outcomes

The study timeline is shown in Figure 1. Study outcomes occurred at any time on or after the first day of the first COVID-19 episode, either (+) or (-). If there were two COVID-19 episodes, the 1^st^ set of outcomes occurred before the 1^st^ day of the second COVID-19 episode, and 2^nd^ set of outcomes occurred on or after the first day of the 2^nd^ COVID-19 episode. If neither a primary nor secondary outcome occurred, such record was censored on the last date the patient was known to be alive and event-free, which per the study design was the date when the study investigator collected EMR data.

**Figure 1.**
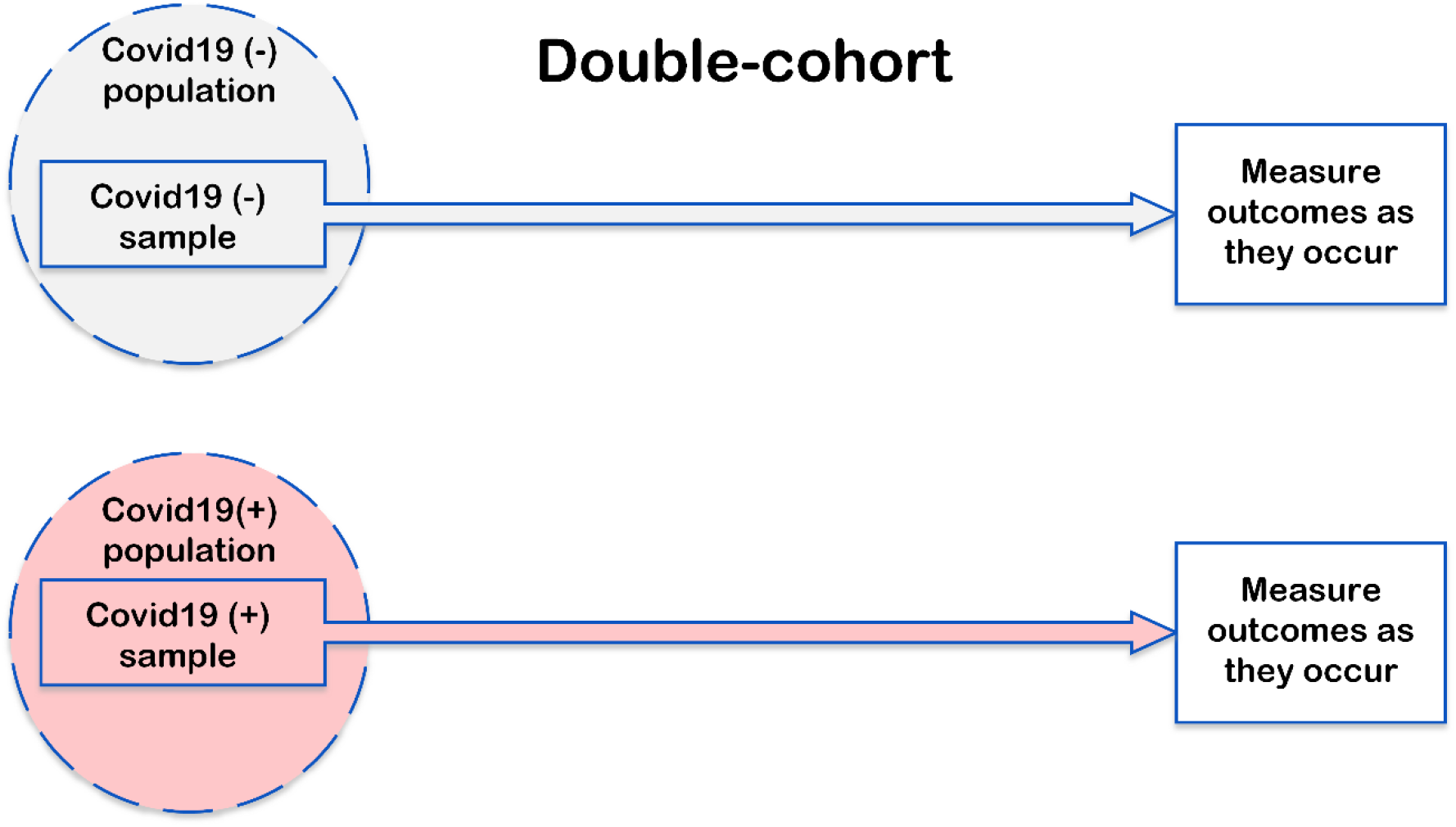
**A.** Study design and analysis.

The primary outcome was defined as a composite of cardiovascular death, acute HF, acute coronary syndrome (STEMI, NSTEMI, or unstable angina), incident stroke or transient ischemic attack, another acute or new cardiovascular outcome prompting healthcare utilization (deep venous thrombosis, pulmonary embolism, pulmonary hypertension, myocarditis, endocarditis, hypertension emergency, or kidney injury[15]), critical care utilization (ICU bed) due to either primary or secondary cardiovascular condition, or development of a life-threatening arrhythmia [sustained ventricular tachycardia(VT)/ventricular fibrillation(VF) or resuscitated sudden cardiac arrest(SCA)], whichever came first. Secondary outcomes included (1) all-cause death and (2) any documented cardiac arrhythmia.

A Mortality & Morbidity Classification Committee (MMCC) was formed and comprised of 3 members (HP, CCM, LGT). The MMCC reviewed and adjudicated outcomes in cases (n= 3) submitted for review by the study investigators. In addition, the MMCC reviewed and adjudicated outcomes in seven randomly selected patient records and confirmed 100% agreement with the adjudication of outcomes performed by the study investigators.

### Definitions of covariates

We collected information on patient demographic characteristics, past medical history and medications, COVID-19 symptoms and treatment, ECG, and echocardiogram measurements, in accordance with the definitions and timeline of COVID-19 episodes. A data dictionary codebook is provided as a Supplement.

Healthy status was documented by regular annual check-ups and an absence of any medical history documented in the EMR. Cardiovascular disease history included at least one of the following: history of hypertension, supraventricular tachycardia (SVT; defined as either atrial fibrillation (AF), flutter, or unspecified SVT), VT/VF, SCA, HF, coronary heart disease (CHD; defined as either prior percutaneous coronary intervention (PCI), coronary artery bypass grafting (CABG), myocardial infarction (MI), documented CHD diagnosis in EMR), dyslipidemia treated by lipid-lowering drugs (LLD), peripheral artery disease (PAD), non-coronary heart disease (defined as either valvular heart disease, congenital heart disease, myocarditis/pericarditis, inherited channelopathy, inherited cardiomyopathy, or pulmonary embolism history), or heart transplant. History of non-coronary atherosclerosis included a history of PAD or carotid artery endarterectomy/stenting. History of cerebrovascular disease included a history of stroke, transient ischemic attack (TIA), or carotid artery endarterectomy/stenting.

Use of medications inhibiting renin-angiotensin-aldosterone system (RAAS) included the use of angiotensin-converting enzyme inhibitors (ACEI), angiotensin receptor blockers (ARB), or angiotensin receptor-neprilysin inhibitor (ARNI). Use of atrioventricular (AV) nodal agents included the use of either beta-blocker (BB), calcium channel blockers (CCB), or class I or class III antiarrhythmic drugs (AAD). Use of anticoagulant or antiplatelet medications included aspirin, P2Y12-inhibitors, warfarin, non-vitamin K antagonist oral anticoagulants (NOAC), heparins, or direct thrombin inhibitors. Use of immunosuppressants included chemotherapy or use of antiretroviral, corticosteroid, hydroxychloroquine, or other immunosuppressant drugs.

Respiratory disease history included a history of asthma, chronic obstructive pulmonary disease (COPD), bronchiectasis, obesity hypoventilation syndrome, sleep apnea, atelectasis, pneumonia, interstitial lung disease, home oxygen use, active or treated tuberculosis, treated or untreated latent tuberculosis infection (LTBI), non-tuberculosis mycobacterium infection or colonization, pneumoconiosis, invasive pulmonary fungal infection, any pulmonary hypertension, or cystic fibrosis. Liver disease history included a history of hepatitis, cirrhosis, hepatocellular carcinoma, metastatic liver cancer, alcoholic liver disease, toxic liver disease, documented liver injury, or gallbladder and biliary tract disease. Kidney disease included a history of glomerulonephritis, chronic kidney disease (CKD), end-stage kidney disease (ESKD), hereditary kidney disorders, obstructive uropathy, or kidney transplant. History of diseases increasing thromboembolism risk was defined as a history of deep venous thrombosis, cardiac septal defect, or coagulopathy. History of diabetes mellitus was defined as a documented prediabetes, diet-controlled, oral hypoglycemic agents-, or insulin-controlled diabetes type I or II. Immunocompromised status was defined as a presence of pregnancy or history of cancer, autoimmune disorder, organ (including bone marrow) transplant, congenital immunodeficiency disorder, use of immunosuppressants, or human immunodeficiency virus (HIV) – positive status. Addiction status was defined as a history of tobacco use (either smoking or chewing), cannabis, alcohol abuse, or illicit drug use. Endocrine disease history was defined as a history of acromegaly, adrenal insufficiency, Cushing’s syndrome, hypo- or hyperthyroidism, parathyroid, or other endocrine disorder treated by an endocrinologist. Blood disease history was defined as a history of anemia, bleeding disorders, hematological malignancies, or hemoglobinopathies. Systemic disease history included amyloidosis, sarcoidosis, hemochromatosis, systemic parasite/infectious disease, or Marfan syndrome.

### Data collection and quality control measures

Data were abstracted from the EMRs by the study investigators. Two investigators (HP and AM) reviewed a random subset of 15 EMR records and confirmed 100% inter-investigator agreement in the definition of exposure, outcomes, and covariates.

### Statistical analysis

Normally distributed continuous variables were summarized as means and standard deviation (SD) and compared using a two-sided *t*-test. Pearson’s χ^2^ test was used to compare categorical variables in patients with two levels of COVID-19 exposure: positive (COVID-19(+) cohort) and negative (COVID-19(-) cohort). COVID-19(+) cohort included patients with COVID-19(+) episodes (either 1^st^ or 2^nd^, or both). COVID-19(-) cohort included patients who had COVID-19(-) episodes only and did not have any COVID-19(+) episodes. The analytical approach is illustrated in Figure 1. The unadjusted Kaplan–Meier survivor functions were plotted for two levels of exposure for the primary and secondary outcomes. We used the log-rank test for the equality of survivor functions across two levels of exposure. Incidence rate and incidence rate difference were calculated to assess the absolute risk and absolute risk difference between two levels of exposure.

To answer a question of whether either asymptomatic or symptomatic SARS-CoV-2 infection is associated with outcomes independently from known COVID-19 risk factors, prevalent CVD, and cardiovascular risk factors, we constructed two Cox proportional hazards models. The proportional-hazards assumption was tested using *stcox* PH-assumptions suite of tests implemented in STATA (StataCorp LP, College Station, TX). Model 1 was adjusted for demographic (age, sex, and race-ethnicity group categories, defined as white non-Hispanic versus non-white or Hispanic) and socioeconomic characteristics (insurance status), and reason for testing (presence or absence of COVID-19 symptoms during the COVID-19 episode). Model 2, in addition to covariates included in model 1, was adjusted for cardiovascular and COVID-19 risk factors (history of CVD, cerebrovascular, liver disease, diabetes mellitus, conditions with an elevated risk of thromboembolism, immunocompromised status, and use of any prescription medication).

In addition, we utilized the causal inference approach and counterfactual analytical framework to investigate the hypothetically causal average treatment effect on the treated (ATET) of COVID-19 exposure on the study outcomes. The ATET estimation has several advantages over the hazard ratio (HR) as an effect estimator. First, the ATET measures the effect in the same time units as the time to outcome instead of in a relative conditional probability.

Second, the models used to estimate ATET are more flexible, as there are no assumptions of linearity and proportional hazards and no risk of model overfitting if too many covariates are included. Nevertheless, ATET estimation requires the assumptions of conditional independence, sufficient overlap, and correct adjustment for censoring. Estimating the ATET requires a significantly weaker version of the conditional independence assumption than estimating the average treatment effect in population (ATE).

We used inverse-probability-weighted (IPW) estimators, using weighted averages of the observed outcome to calculate the potential-outcome means (POMs) and ATET. The IPW estimators were implemented in a three-step approach. First, we estimated the parameters of a treatment-assignment model (predicting probabilities of a subject to be included in a COVID-19(+) or COVID-19(-) cohort) and computed the component of the estimated weights that accounts for data missing because each subject was only observed after receiving one of the possible treatment levels, either COVID-19(+) or COVID-19(-). The model was conditioned for 31 covariates, including demographic and socioeconomic characteristics (age, sex, race, and ethnicity, health insurance status), the reason for COVID-19 testing, medical history of CVD, cerebrovascular, respiratory, kidney, liver, blood, systemic, endocrine disease, diabetes mellitus, addiction, conditions with immunocompromised and thromboembolic risk, use of prescription medications (including RAAS-blocking drugs, AV-nodal agents, antiplatelet or anticoagulant, and immunosuppressants), and the presence and type of COVID-19 symptoms (fever, fatigue, runny nose, headache, muscle and body aches, cough, shortness of breath, ageusia or anosmia, and nausea). Next, we estimated the parameters of a time-to-censoring model and computed the component of estimated weights that accounts for data lost to censoring. In this retrospective study, the censoring time was determined by a random day when a study investigator collected EMR data unless a patient utilized healthcare and experienced potential outcome. Thus, we assumed that the time to censoring was random. We conditioned the model predicting time-to-censoring for the same 31 covariates as described above for the model predicting treatment assignment. At the final third step, we used both estimated weights to compute weighted averages of the outcomes for the COVID-19(+) cohort.

We conducted balance checks for the treatment-assignment model. We tested an overlap assumption that each study participant has a sufficient positive probability of being assigned to each treatment level. These checks depend only on the estimated probabilities of COVID-19(+) cohort assignment and are not affected by the censoring of the outcome. We observed (Supplemental Table 1) that the weighted standardized differences are much closer to 0 than the raw standardized differences, and the weighted variance ratios are much closer to 1 than the raw variance ratios. Therefore, we concluded that the model-based treatment weights balanced the covariates. We conducted a formal test of the hypothesis that the weighted constructed from the treatment-assignment model balances the covariates. We observed that we do not reject the null hypothesis that the treatment-assignment model is well specified (*p*=0.965; Figure 2). Thus, we used this model to look for evidence that the overlap condition is violated. Figure 2 showed that the densities for the probability to be included in the COVID-19(+) cohort were evenly distributed and showed sufficient overlap, and the maximum probability to be included in either COVID-19(+) or COVID-19(-) cohort was sufficiently less than 1. However, the densities for the probability to be included in the Covid19(-) cohort violated the overlap assumption, indicating that there were unmeasured patients’ characteristics that increased the probability for a patient to belong to COVID-19(-) cohort, likely because many of these patients underwent unrelated to COVID-19 medical procedures, and were tested for COVID-19 as a part of hospital precautions. Therefore, we reported only ATET estimators and not ATE estimators.

**Table 1.** Baseline clinical and demographic characteristics in Covid19 (+) and (-) cohorts

| Characteristic | Covid19(+) cohort<br>(n=319) | Covid19(-) cohort<br>(n=1,036) | p-value |
| --- | --- | --- | --- |
| **Age(SD), y** | **46.7(18.5)** | **49.4(21.1)** | **0.032** |
| Female, n(%) | 186(58.3) | 584(56.4) | 0.541 |
| **White non-Hispanic, n(%)** | **158(49.5)** | **819(79.1)** | **<0.0001** |
| **Insured, n(%)** | **226(70.6)** | **846(81.7)** | **<0.0001** |
| Healthy, n(%) | 61(19.1) | 212(20.5) | 0.602 |
| Cardiovascular disease Hx, n(%) | 129(40.4) | 434(41.9) | 0.645 |
| Hypertension Hx, n(%) | 86(27.0) | 321(31.0) | 0.170 |
| **SVT (including AF) Hx, n(%)** | **15(4.7)** | **83(8.0)** | **0.046** |
| VT or SCA Hx, n(%) | 3(0.9) | 8(0.8) | 0.770 |
| Heart failure Hx, n(%) | 14(4.4) | 64(6.2) | 0.230 |
| **Any CHD Hx, n(%)** | **12(3.8)** | **87(8.4)** | **0.005** |
| Dyslipidemia on LLD, n(%) | 73(22.9) | 276(26.6) | 0.180 |
| **Noncoronary atherosclerosis Hx, n(%)** | **2(0.6)** | **32(3.1)** | **0.014** |
| Noncoronary heart disease, n(%) | 13(4.1) | 58(5.6) | 0.286 |
| Cerebrovascular disease Hx, n(%) | 16(5.0) | 57(5.5) | 0.737 |
| Respiratory disease Hx, n(%) | 98(30.7) | 264(25.5) | 0.064 |
| **Liver disease Hx, n(%)** | **38(11.9)** | **75(7.2)** | **0.008** |
| Kidney disease Hx, n(%) | 32(10.0) | 114(11.0) | 0.624 |
| Thromboembolism risk Hx, n(%) | 7(2.2) | 41(4.0) | 0.136 |
| **Diabetes mellitus Hx, n(%)** | **81(25.4)** | **196(18.9)** | **0.012** |
| Immunocompromised Hx, n(%) | 53(16.6) | 204(19.7) | 0.220 |
| Smoking & Addiction Hx, n(%) | 68(21.3) | 225(21.7) | 0.879 |
| **Endocrine disease Hx, n(%)** | **31(9.7)** | **147(14.2)** | **0.039** |
| Blood disease Hx, n(%) | 56(17.6) | 163(15.7) | 0.440 |
| Systemic disease Hx, n(%) | 4(1.3) | 13(1.3) | 0.999 |
| **On any Rx medication, n(%)** | **177(55.5)** | **504(48.6)** | **0.033** |
| RAAS medication use, n(%) | 47(14.7) | 153(14.8) | 0.988 |
| AV nodal agents use, n(%) | 41(12.9) | 172(16.6) | 0.108 |
| Anticoagulant/antiplatelet use, n(%) | 51(16.0) | 213(20.6) | 0.071 |
| Immunosuppressant use, n(%) | 31(9.7) | 112(10.8) | 0.579 |

SVT=supraventricular tachycardia; AF=atrial fibrillation; VT=ventricular tachycardia;
SCA=sudden cardiac arrest; Hx=history; CHD=coronary heart disease; AV=atrioventricular;
Rx=prescribed.

**Figure 2.**
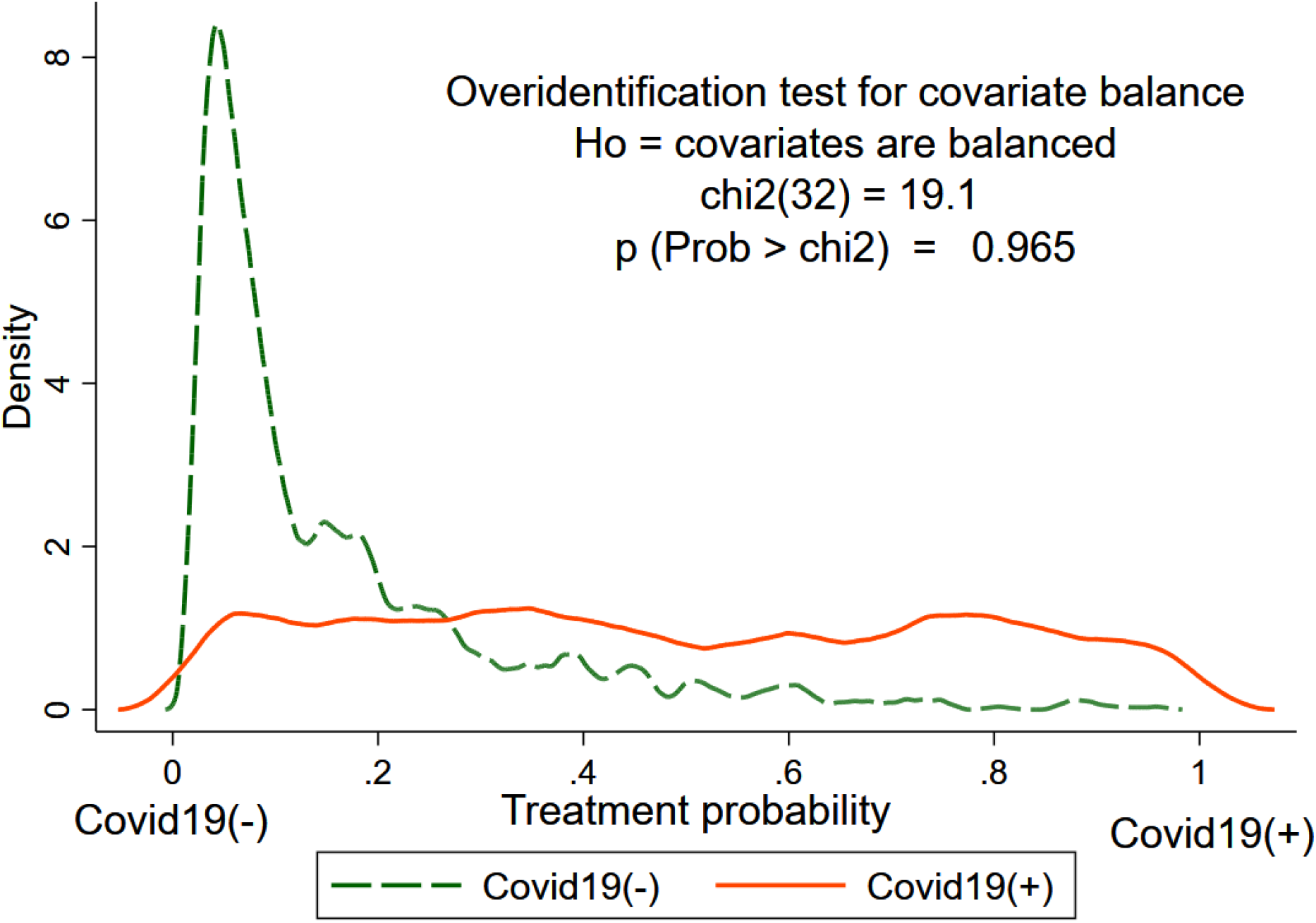
An overlap plot for the estimated densities of the probabilities of being from COVID-19(-) cohort (score=0) or COVID-19(+) cohort (score=1), conditioned for 32 covariates included in inverse-probability-weighted (IPW) model.

Statistical analyses were performed using STATA MP 17 (StataCorp LP, College Station, TX). A *p-*value < 0.05 was considered statistically significant. STATA do files are available at https://github.com/Tereshchenkolab/statistics.

## Results

### Study population

Between March 1^st^, 2020, and September 13^th^, 2020, OHSU performed 99,711 COVID-19 tests for 65,585 individuals. The study investigators included a random sample of 1355 eligible OHSU patient records. Clinical characteristics of the patient population are presented in Table 1.

COVID-19(+) patients were younger, more likely to be non-white or Hispanic, and less likely to be insured than COVID-19(-) patients. Furthermore, COVID-19(+) patients were more likely to have been prescribed medication and have a history of liver disease and/or diabetes mellitus (Table 1). There was no difference in CVD history between the two exposure cohorts.

### Exposure: COVID-19 manifestation and timeline between COVID-19 episodes

There was a significant difference in the reasons for COVID-19 testing between two cohorts: COVID-19(+) cohort (n=319) patients were twice as likely to be symptomatic than COVID-19(-) cohort (n=1036) patients (Table 2). All COVID-19(+) episodes were confirmed by a PCR test, whereas 17 COVID-19(-) episodes (0.02%) were detected by an antibody test.

**Table 2.** Covid19 exposure characteristics

| Characteristic of Covid19 episode | Covid19(+) cohort<br>(n=319) | Covid19(-) cohort<br>(n=1,036) | p-value |
| --- | --- | --- | --- |
| Reason for testing: symptomatic patient, n(%) | 255(79.9) | 383(36.8) | <0.0001 |
| Fever, chills, n(%) | 133(41.7) | 127(12.3) | <0.0001 |
| Weakness, fatigue, n(%) | 52(16.3) | 81(7.8) | <0.0001 |
| Muscle and body aches, n(%) | 84(26.3) | 78(7.5) | <0.0001 |
| Runny nose, congestion, sore throat, n(%) | 100(31.4) | 151(14.6) | <0.0001 |
| Cough, n(%) | 147(46.1) | 151(14.6) | <0.0001 |
| Shortness of breath, difficulty breathing, n(%) | 75(23.5) | 99(9.6) | <0.0001 |
| Loss of taste(ageusia) or smell(anosmia), n(%) | 54(16.9) | 10(1.0) | <0.0001 |
| Nausea, vomiting, n(%) | 42(13.2) | 60(5.8) | <0.0001 |
| Anorexia, n(%) | 5(1.6) | 2(0.2) | 0.003 |
| Diarrhea, n(%) | 32(10.0) | 47(4.5) | <0.0001 |
| Headache, n(%) | 90(28.2) | 119(11.5) | <0.0001 |
| Confusion, n(%) | 4(1.3) | 9(0.9) | 0.537 |
| Pain or pressure in the chest, n(%) | 22(6.9) | 28(2.7) | 0.001 |
| Other symptoms, n(%) | 19(6.0) | 30(2.9) | 0.010 |

All known COVID-19 symptoms were more frequently observed in COVID-19(+) patients (Table 2). The most frequent COVID-19 symptoms were cough and fever, observed in > 40% of patients. Runny nose, headache, muscle and body aches, and shortness of breath were present in > 20% of patients. Loss of taste (ageusia) or smell (anosmia) was present in approximately 17% of patients, and fatigue in approximately 16% of patients. Nausea, vomiting, diarrhea, anorexia, and chest pain were less frequent than other symptoms. In addition to the symptoms listed in Table 2, COVID-19(+) patients also suffered from abdominal pain, ear pain, dizziness, vertigo, hemoptysis, and dark stool. Notably, 20% of COVID-19(+) patients were asymptomatic.

Most patients had a single COVID-19 episode, either positive or negative. Four out of 319 patients (1.25%) had reinfection that occurred 58.5±23.6 days (range 33-89 days) after the first COVID-19(+) episode. Thirty-one patients experienced COVID-19(+) episode 94.5±61.3 days (range 15-247 days) after COVID-19(-) episode. Thirty-five patients had COVID-19(-) episode 145.5±90.9 days (range 36-371 days) after COVID-19(+) episode. Because only a small number of patients experienced reinfection or two different types of COVID-19 episodes, we were precluded from completing a meaningful crossover analysis.

### Primary composite outcome analysis

During a median of 178 days at risk, the primary composite outcome was observed in a total of 103 patients, 38 (12%) of whom were from the COVID-19(+) cohort, and 65 (6%) were from COVID-19(-) cohort (*p*=0.001). Acute HF was diagnosed in 7, SCA/VF in 2, STEMI in 1, NSTEMI in 5, incident stroke in 5, endocarditis in 3, DVT/pulmonary embolism in 6, acute kidney dysfunction in 24, critical care utilization due to an acute primary or secondary cardiovascular condition in 25, and cardiovascular death in 25. Those who developed primary outcome were more likely to have greater severity of COVID-19 (Supplemental Table 2). However, only 26% of them were hospitalized because of COVID-19, and only 13% utilized ICU beds.

#### Absolute (attributable) risk of the primary composite outcome

Among COVID-19(+) cohort participants, the incidence rate of the primary outcome was higher (178.6 per 1,000 person-years of follow-up) than among COVID-19(-) cohort participants (149.2 per 1,000 person-years of follow-up). However, the incidence rate difference in the primary outcome between the two levels of exposure did not reach statistical significance (29.4; 95% CI from -38.04 to 96.8 per 1,000 person-years of follow-up; *p*=0.379).

#### The relative risk of COVID-19(+) exposure as compared to COVID-19(-) exposure

In unadjusted survival analysis, COVID-19(+) patients had a significantly higher probability of developing the primary composite outcome than COVID-19(-) patients (Figure 3). In unadjusted Cox regression analysis, COVID-19(+) exposure was associated with a more than 50% higher risk of the primary outcome (Table 3). After adjustment for demographic characteristics, health insurance status, and reason for COVID-19 testing (model 1), COVID-19 infection remained associated with the primary outcome. However, the association attenuated after additional adjustment for prevalent CVD, cardiovascular, and COVID-19 risk factors in model 2. Proportional-hazards assumption was confirmed for all Cox regression models with the primary composite outcome.

**Table 3.** Association of COVID-19 exposure with the study outcomes in Cox regression models.

| Model | Composite primary outcome |  | All-cause death |  |
| --- | --- | --- | --- | --- |
|  | HR (95% CI) | p-value | HR (95%CI) | p-value |
| Unadjusted | 1.54(1.02-2.34) | 0.042 | 1.21(0.56-2.63) | 0.631 |
| Model 1 | 1.71(1.06-2.78) | 0.029 | 1.27(0.52-3.12) | 0.600 |
| Model 2 | 1.47(0.90-2.38) | 0.122 | 1.08(0.44-2.65) | 0.874 |

**Figure 3.**
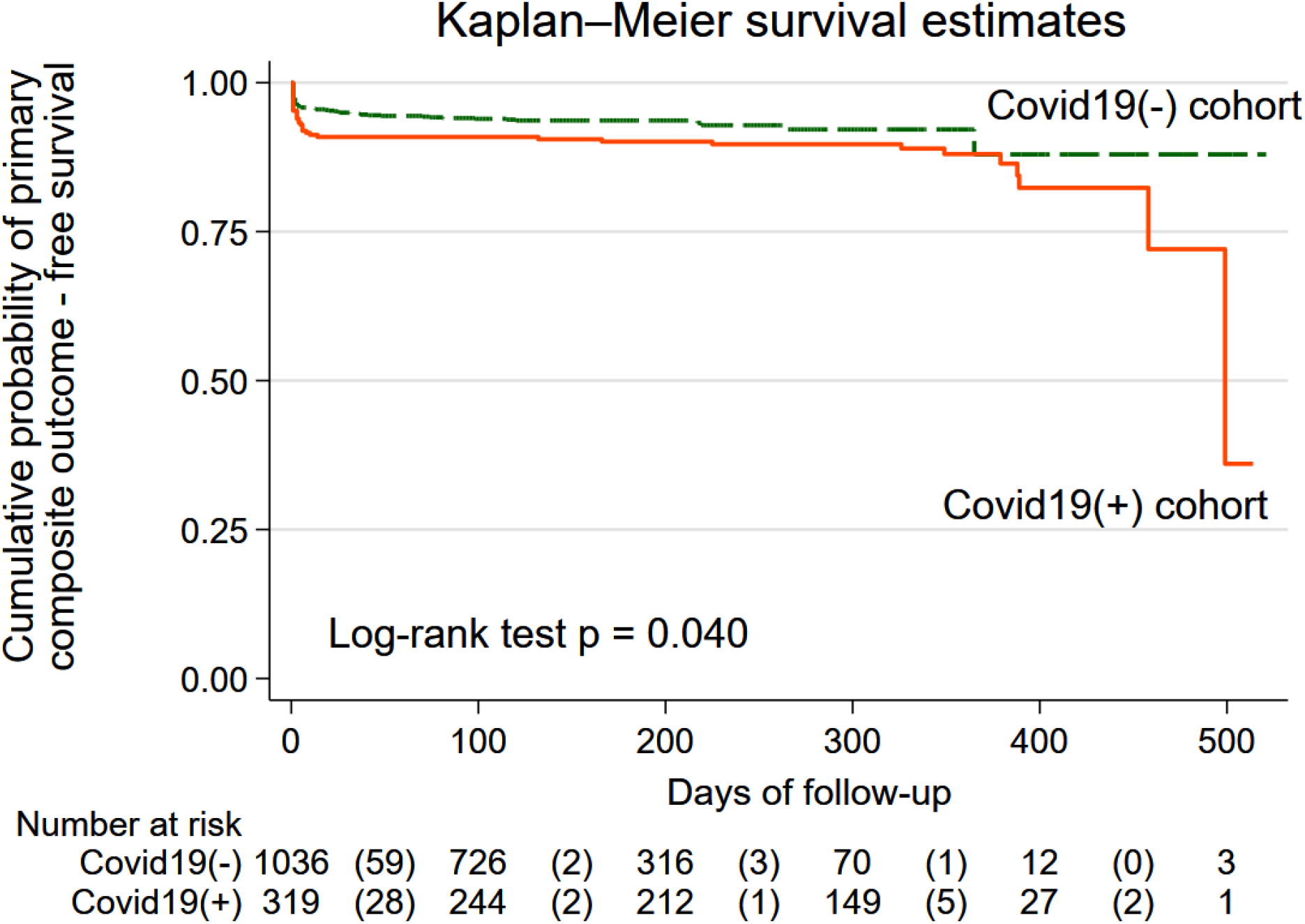
The estimated unadjusted Kaplan-Meier survivor functions for the primary composite outcome in COVID-19(+) (solid orange line) and COVID-19(-) (green dashed line) cohorts. The table below the graph shows the number at risk in each group at every 100 days of follow-up. The number of primary composite outcome events at every 100 days of follow-up is shown in parenthesis.

#### Causal inference analysis

In causal inference analysis (Table 4), in the COVID-19(+) cohort, the average time to the primary composite outcome was estimated to be 163.8 days or approximately 5.4 months *more* than when everyone in the COVID-19(+) cohort was COVID-19(-). The estimated average time to the primary composite outcome when all in the COVID-19(+) cohort were COVID-19(-) was 148.5 days or approximately 4.9 months.

**Table 4.**
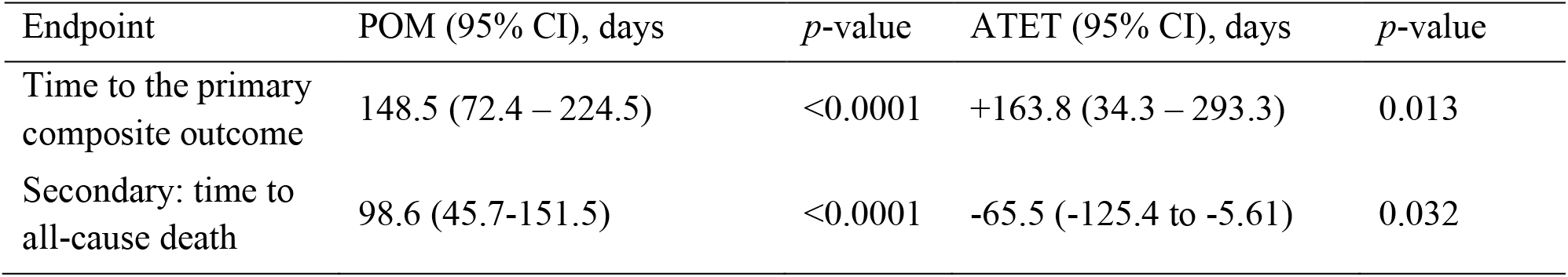
The average treatment effect on the treated (ATET) for COVID-19(+) versus COVID-19(-), and the potential-outcome means (POMs) estimates for COVID-19(-) cohort participants.

### Secondary outcome: all-cause mortality analyses

During median 190 days at risk, there were 32 all-cause deaths: 10 deaths in the COVID-19(+) cohort and 22 in the COVID-19(-) cohort. Among COVID-19(+) cohort participants, the incidence rate of the all-cause death was 41.6 per 1,000 person-years of follow-up, in comparison to 45.1 per 1,000 person-years of follow-up among COVID-19(-) cohort participants. There was no statistically significant incidence rate difference (−3.4; 95% CI from - 35.4 to 28.5 per 1,000 person-years of follow-up; *p*=0.855) in the all-cause death between two cohorts.

In Kaplan-Meier survival analysis, there were no differences in all-cause mortality between COVID-19(-) and COVID-19(+) cohorts (Figure 4). In unadjusted Cox regression analysis, COVID-19(+) exposure was associated with non-significant risk (Table 3). Notably, the proportional-hazards assumption was violated for all Cox regression models with the all-cause death outcome.

**Figure 4.**
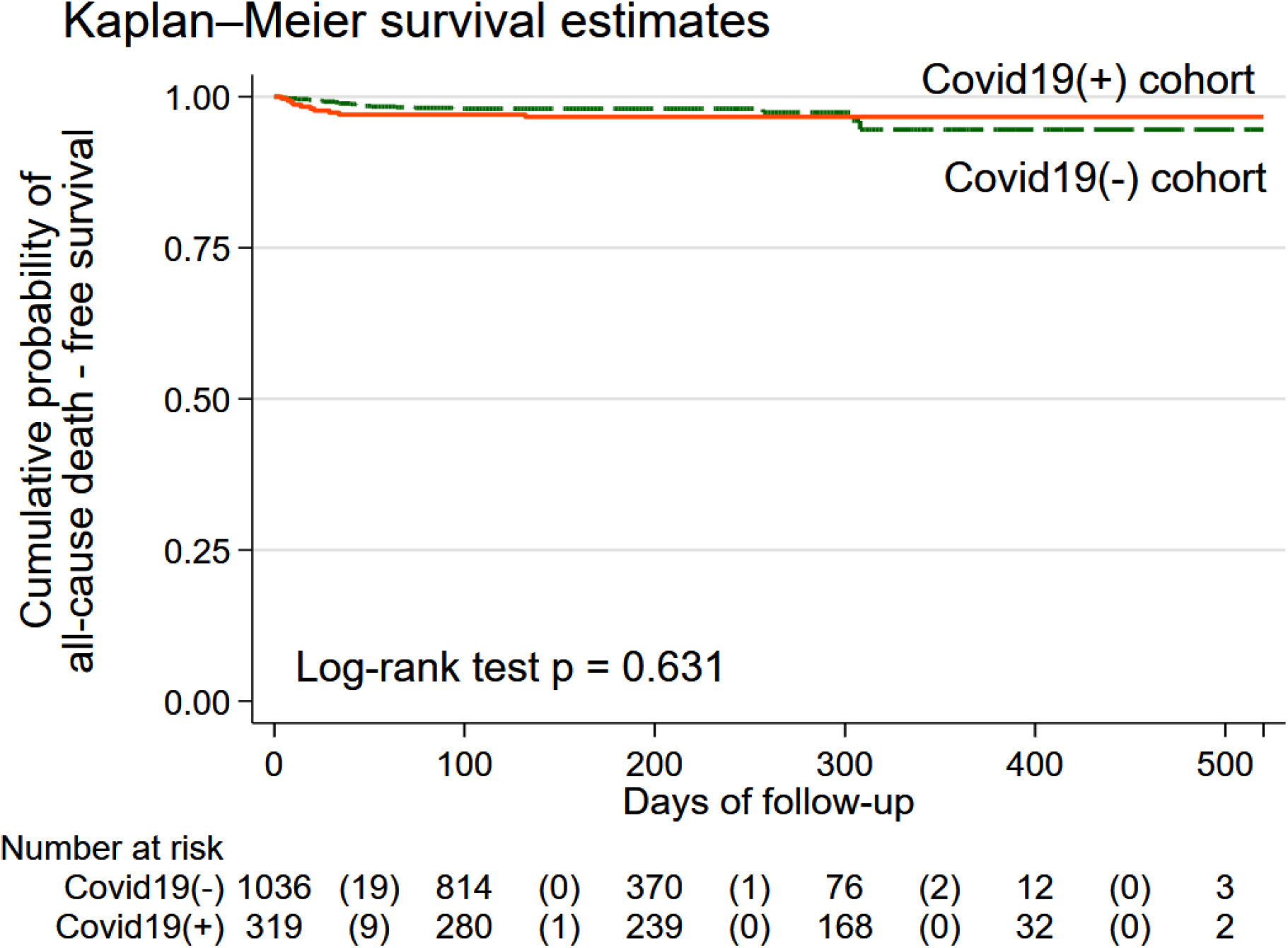
The estimated unadjusted Kaplan-Meier survivor functions for all-cause mortality in COVID-19(+) (solid orange line) and COVID-19(-) (green dashed line) cohorts. The table below the graph shows the number at risk in each group at every 100 days of follow-up. The number of primary composite outcome events at every 100 days of follow-up is shown in parenthesis.

Importantly, causal inference analysis using IPW estimators, conditioned for 31 covariates as described above, showed that for every COVID-19(+) patient, the average time to all-cause death was estimated to be 65.5 days *less* than when all these patients were COVID-19(-). The estimated average time to all-cause death when all these patients were COVID-19(-) was 98.6 days (Table 4).

Another pre-specified secondary outcome, documented cardiac arrhythmia, was recorded in only 10 study participants: 4 in the COVID-19(+) cohort and 9 in the COVID-19(-) cohort. Therefore, we did not conduct survival analyses due to a small number of documented cardiac arrhythmia outcomes.

## Discussion

In this retrospective double-cohort study, after rigorous adjustment for demographic and socioeconomic characteristics, reasons for COVID-19 testing, acute COVID-19 symptoms, medical history, risk factors of both COVID-19 and CVD, and use of medications, we found that either symptomatic or asymptomatic SARS-CoV-2 infection was associated with increased risk of late cardiovascular outcomes, occurring at least 30 days (on average 10 months) after SARS-CoV-2 infection. Importantly, we demonstrated the effect of COVID-19 infection on cardiovascular events, regardless of initial presenting COVID-19 symptoms. This finding highlights the importance of COVID-19 prevention and suggests that careful follow-up might be needed for any patient who experienced SARS-CoV-2 infection, either symptomatic or asymptomatic, to monitor for late cardiovascular events. The second important finding of our study was the causal effect of either symptomatic or asymptomatic SARS-CoV-2 infection on all-cause death occurring during the post-acute or late COVID-19 period.

There is both pathophysiological basis and clinical evidence of significant cardiovascular risk following COVID-19.[2] Several recent studies confirmed the risks of long-term cardiovascular consequences of COVID-19 while showing a wide range of an estimated disease burden.[8, 16, 17] Several small cardiac magnetic resonance (CMR) case-control studies of patients who recovered from mild or moderate COVID-19 showed a high prevalence (71-78%) of CMR abnormalities.[8, 16] However, other small (n=58) case-control studies reported relatively low (21%) prevalence of CMR abnormalities 6 months after moderate-to-severe COVID-19.[18] Furthermore, a well-matched case-control (n=75) 6-month post-COVID-19 CMR study concluded that there were no differences in the left ventricular (LV) structure, function, and scar burden between cases and controls.[19] Cohort studies have advantages over case-control studies as all individuals are derived from the same study population, and there is no uncertainty regarding the time of exposure preceding the outcome when establishing a cause and effect relationship. Our double-cohort study was the first to estimate both absolute (attributable) and relative risks of SARS-CoV-2 infection for the development of late cardiovascular outcomes.

Relatively few longitudinal COVID-19 cardiovascular studies have been reported up to date. Chaturvedi et al. in a prospective echocardiographic study of hospitalized, symptomatic COVID-19 patients, reported a decline in both left and right heart function 3 months after hospital discharge.[20] Preliminary findings of the Prospective longitudinal study Capturing MultiORgan Effects of COVID-19 (C-MORE)[21] showed that more than half of the patients experienced symptoms at 6 months post-COVID-19, limiting their ability to exercise. C-MORE investigators also noted a dissociation between symptoms and objective measures of cardiovascular health.[21] Our study did not ascertain the duration of the COVID-19 symptoms,[22] which should be further studied in future prospective studies.

Our study contributed to the growing body of knowledge showing the cardiovascular implications of SARS-CoV-2 infection regardless of its symptoms. Consistently with our findings, a large study of the English National Immunisation (NIMS) Database of COVID-19 vaccination, using self-controlled case series methodology, showed that SARS-CoV-2 infection was associated with a substantial increase in the risk of hospitalization or death from myocarditis, pericarditis, and cardiac arrhythmia.[23] The distinct mechanism of SARS-CoV-2 infection includes ACE2 downregulation, diminishing the protective, anti-inflammatory role of ACE2, and, thus, facilitating myocardial injury and fibrosis as the virus’s long-term sequelae.[24] Very few studies have investigated the post-mortem cardiac pathology of patients who died with severe COVID-19. Available data indicates a modest histopathological involvement of the heart and mostly nonspecific findings of coronary atherosclerosis and left ventricular hypertrophy.[25] Development of the pulmonary vascular thrombosis is a frequently observed complication of COVID-19 pneumonia.[26] Frequently observed nonspecific cardiac pathology in COVID-19 highlights the importance of appropriate control in study design assessing cardiovascular risks of SARS-CoV-2 infection. Any pandemic or epidemic (regardless of the type of a pathogen) might be associated with increased cardiovascular mortality.[27] The double-cohort design used in this study allowed us to demonstrate the causal nature of SARS-CoV-2 infection with cardiovascular events.

Our COVID-19(-) and COVID-19(+) cohorts had a similar absolute number of all-cause deaths, consistent with the notion about indirect consequences of the pandemic, as the healthcare systems redistributed resources towards COVID-19 patients, while the standards of health care delivery have been reduced.[28] Using a counterfactual analytical framework, we showed that all-cause death in the COVID-19(+) cohort occurred by 2 months sooner than if all these patients did not experience SARS-CoV-2 infection. Importantly, we conditioned for 31 covariates, including a comprehensive list of demographic and socioeconomic characteristics, medical history and treatment, and COVID-19 symptoms. Our finding of a causal effect of either symptomatic or asymptomatic SARS-CoV-2 infection on all-cause mortality supports previous reports linking excess all-cause mortality during the pandemic with SARS-CoV-2 infection.[14] We observed a low reinfection rate, consistent with other studies.[29]

We found that COVID-19(+) patients were more likely to be non-white or Hispanic and less likely to be insured than COVID-19(-) patients. Furthermore, other studies have shown that racial and ethnic minority groups have a significantly higher risk of COVID-19 positivity and that socioeconomic determinants were strongly associated with outcomes.[30] [27, 28] This recurrent disproportionality suggests that health inequities and socioeconomic determinants play a significant role in the ongoing COVID-19 pandemic and that interventions should be aimed at mitigating these negative impacts.

### Limitations

Important limitations of the study need to be considered. As in any retrospective cohort study, investigators had no control over the quality and completeness of the available EMR data. The likelihood of unobserved and unmeasured confounding cannot be eliminated entirely, as an observational study is susceptible to confounding bias. Two cohorts assembled from the different COVDI-19(+) and COVID-19(-) populations may differ in multiple important ways that influenced the outcomes. We cannot completely rule out the violation of the conditional independence assumption. In our observational study, the treatment (SARS-CoV-2 infection exposure) was not randomly assigned so potential outcomes are not independent of the exposure. We assumed that after conditioning on the covariates, the treatment assignment was as good as random. Nevertheless, we cannot be 100% sure that we observed, measured, and conditioned on enough covariates. We also note that the study was conducted in a single healthcare system. Validation of the study findings in alternative populations will increase the chances that the observed association is causal.

## Data Availability

All data produced in the present study are available upon reasonable request to the authors

## Acknowledgment

We would like to thank the following study investigators for their help with data collection: Hannah Kim, Isabelle Nguyen, Linh Nhat Taylor, Brianna Pickering, Maya Herzig, Olivia Glatt, Tahmina Karimyar, and Timothy Kang.

---

**Supplemental Table 1.** Covariate balance summary.

| Covariate | Standardized differences |  | Variance ratio |  |
| --- | --- | --- | --- | --- |
|  | Raw | Weighted | Raw | Weighted |
| Age | -0.133 | -0.016 | 0.771 | 0.921 |
| Sex | -0.039 | -0.097 | 0.991 | 0.977 |
| White non-Hispanic | -0.647 | -0.138 | 1.513 | 1.017 |
| Health insurance | 0.256 | -0.134 | 1.513 | 1.017 |
| Reason for Covid19 testing | 0.970 | -0.017 | 0.690 | 1.027 |
| CVD History | -0.029 | -0.014 | 0.992 | 0.995 |
| Respiratory History | 0.117 | -0.113 | 1.123 | 0.923 |
| Cerebrovascular disease history | -0.022 | 0.069 | 0.918 | 1.366 |
| Kidney disease history | -0.032 | 0.024 | 0.924 | 1.069 |
| Liver disease history | 0.159 | 0.115 | 1.566 | 1.359 |
| Diabetes history | 0.156 | -0.021 | 1.238 | 0.978 |
| Conditions with thromboembolic risk | -0.102 | 0.035 | 0.566 | 1.281 |
| Immunocompromised status history | -0.080 | 0.071 | 0.878 | 1.147 |
| Addiction history | -0.010 | -0.269 | 0.989 | 0.756 |
| Other endocrine disease history | -0.138 | -0.016 | 0.722 | 0.957 |
| Blood disease history | 0.049 | 0.020 | 1.094 | 1.035 |
| Systemic disease history | -0.00005 | 0.017 | 1.001 | 1.169 |
| No prescription medications | 0.137 | 0.060 | 0.991 | 0.990 |
| RAAS-blockers | -0.001 | 0.004 | 1.000 | 1.009 |
| AV-nodal agents | -0.106 | 0.015 | 0.811 | 1.033 |
| Anticoagulants/antiplatelet agents | -0.118 | -0.028 | 0.824 | 0.950 |
| Immunosuppressors | -0.036 | -0.018 | 0.911 | 0.952 |
| Covid19 symptom fever | 0.702 | 0.008 | 2.265 | 1.003 |
| Covid19 symptom fatigue | 0.262 | 0.016 | 1.897 | 1.031 |
| Covid19 symptom muscle aches | 0.517 | -0.005 | 2.792 | 0.994 |
| Covid19 symptom running nose | 0.406 | 0.085 | 1.732 | 1.080 |
| Covid19 symptom cough | 0.729 | -0.016 | 2.000 | 0.998 |
| Covid19 symptom shortness of breath | 0.382 | -0.126 | 2.085 | 0.872 |
| Covid19 symptom ageusia/anosmia | 0.582 | -0.144 | 14.74 | 0.802 |
| Covid19 symptom nausea | 0.253 | 0.087 | 2.100 | 1.232 |
| Covid19 symptom headache | 0.428 | 0.006 | 2.000 | 1.006 |

**Supplemental Table 2.**
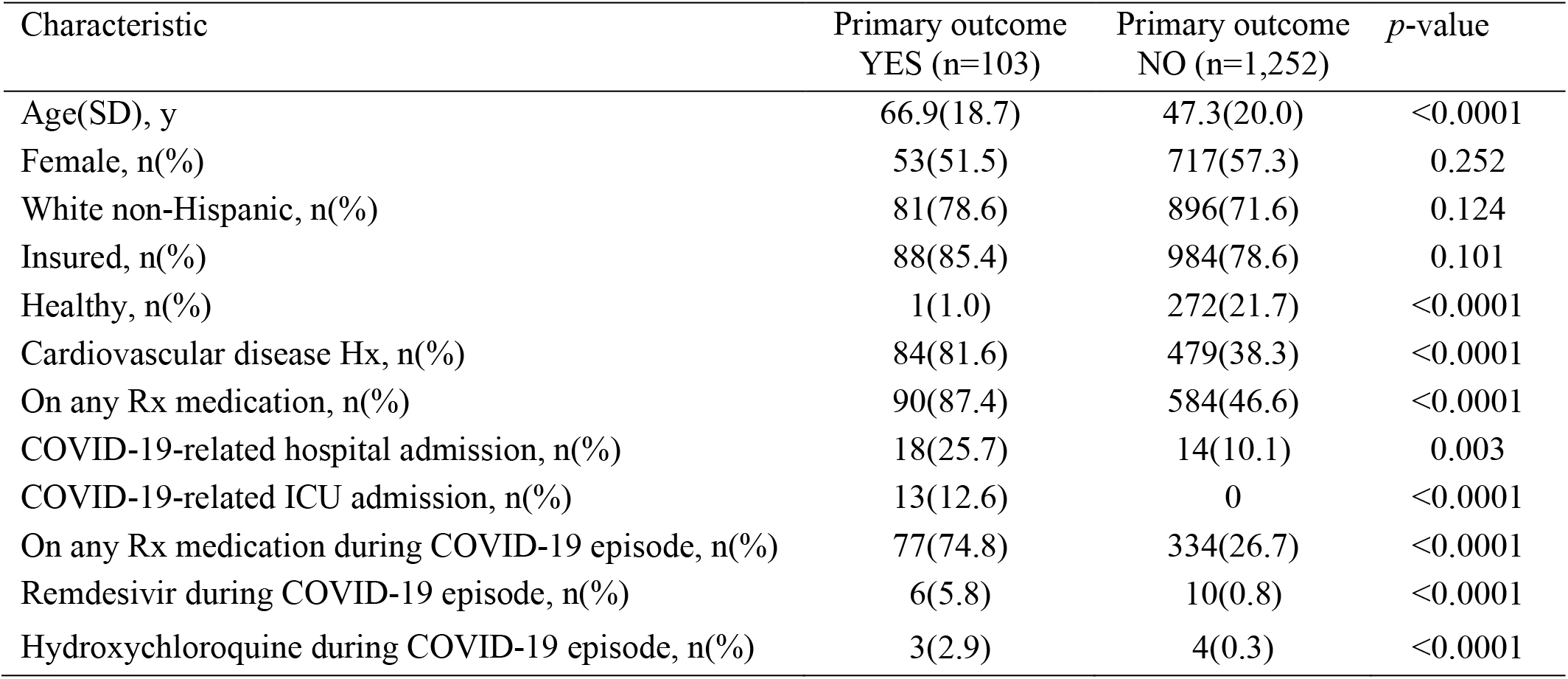
Comparison of patients characteristics by primary outcome.

